# Guidance for triangulating data and estimates of HIV prevalence among pregnant women and coverage of PMTCT using the Spectrum AIDS Impact Module

**DOI:** 10.1101/2024.05.29.24306862

**Authors:** Magdalene K. Walters, Eline L. Korenromp, Anna Yakusik, Ian Wanyeki, André Kaboré, Arthur Poimouribou, Célestine Ki, Coumbo Dao, Paul Bambara, Salam Derme, Théophile Ouedraogo, Kai Hon Tang, Marie-Claude Boily, Mary Mahy, Jeffrey W. Imai-Eaton

**Author notes:** **Correspondence to:** Magdalene Walters, Imperial College London, St. Mary’s Hospital Campus, Norfolk Place, London W2 1P, UK.

## Abstract

**Background:** Most countries use the Spectrum AIDS Impact Module (Spectrum-AIM), antenatal care routine HIV testing, and antiretroviral treatment data to estimate HIV prevalence among pregnant women. Non-representative programme data may lead to inaccurate estimates HIV prevalence and treatment coverage for pregnant women.

**Setting:** 154 locations in 126 countries.

**Methods:** Using 2023 UNAIDS HIV estimates, we calculated three ratios: (1) HIV prevalence among pregnant women to all women 15-49y (prevalence), (2) ART coverage before pregnancy to women 15-49y ART coverage (ART pre-pregnancy), and (3) ART coverage at delivery to women 15-49y ART coverage (PMTCT coverage). We developed an algorithm to identify and adjust inconsistent results within regional ranges in Spectrum-AIM, illustrated using Burkina Faso’s estimates.

**Results:** In 2022, the mean regional ratio of prevalence among pregnant women to all women ranged from 0.68 to 0.95. ART coverage pre-pregnancy ranged by region from 0.40 to 1.22 times ART coverage among all women. Mean regional PMTCT coverage ratios ranged from 0.85 to 1.51. The prevalence ratio in Burkina Faso was 1.59, above the typical range 0.62-1.04 in western and central Africa. Antenatal clinics reported more PMTCT recipients than estimated HIV-positive pregnant women from 2015 to 2019. We adjusted inputted PMTCT programme data to enable consistency of HIV prevalence among pregnant women from programmatic routine HIV testing at antenatal clinics with values typical for Western and central Africa.

**Conclusion:** These ratios offer Spectrum-AIM users a tool to gauge the consistency of their HIV prevalence and treatment coverage estimates among pregnant women with other countries in the region.

## Introduction

Estimates of HIV prevalence among pregnant women determine need for and coverage of antiretrovirals for prevention of maternal to child transmission (PMTCT), a key input to estimating paediatric HIV infections and monitoring progress towards eliminating mother-to-child HIV transmission.^1–4^ Most countries estimate the number of pregnant women living with HIV (PWLHIV) and PMTCT coverage using the AIDS Impact Module in Spectrum (Spectrum-AIM). Comparing related model outcomes and typical patterns across locations can reveal inconsistencies in input data or model assumptions that may result in inaccurate estimates.

Country-specific estimates of HIV prevalence among pregnant women are the result of a multi-step modelling process in Spectrum-AIM. The number of PWLHIV is calculated by multiplying the age-specific number of women living with HIV (WLHIV) in reproductive ages (15 to 49 years [15-49y]), age-specific fertility rates (by 5-year age group), and the relative fertility of WLHIV relative to women without HIV (fertility rate ratio [FRR]). Age-specific HIV prevalence is derived by fitting a mathematical model to local HIV surveillance data,^5–7^ including HIV prevalence from household surveys, prevalence among pregnant women attending antenatal clinics (ANC), prevalence surveys among key populations, new HIV diagnoses, or AIDS-related deaths. Age-specific fertility rates are obtained from the United Nations World Population Prospects.^8^ Relative fertility of WLHIV compared to women without HIV by age, treatment-status, and local effects are estimated from national household surveys in African countries that measure HIV status and collect birth histories.^8,9^ Inconsistencies in population prevalence, fertility rates, or relative fertility among WLHIV may yield inaccurate estimates of HIV prevalence and treatment coverage among pregnant women within Spectrum-AIM.

Quality of information about HIV among pregnant women varies by region and epidemic type. In most African countries with high HIV prevalence, ANC attendance and routine HIV testing at ANC (ANC-RT) is nearly universal. Universal HIV testing at ANC provides direct measures of HIV prevalence among pregnant women. Spectrum-AIM calculates a ‘local adjustment factor’ (LAF) to calibrate the modelled prevalence among pregnant women to HIV prevalence measured from national ANC-RT. The default LAF value of 1.0 indicates that the fertility of WLHIV is consistent with the default HIV FRR estimated from household survey data. In the 2023 UNAIDS estimates, most sub-Saharan African (SSA) locations (n=36/43) calibrated the LAF to ANC-RT data.^10,11^

In countries with low HIV prevalence and transmission primarily among key populations and their partners, mostly outside SSA, ANC HIV testing coverage varies and is often prioritised among higher risk women or higher burden locations. Consequently, HIV prevalence from ANC-RT in these locations does not represent prevalence among all pregnant women and cannot be used to calibrate the LAF.^12–18^ Data are limited about WLHIV’s fertility relative to women without HIV in these settings, and fertility may be more heterogeneous given the greater concentration of HIV among populations such as sex workers, partners of men who have sex with men, and people who inject drugs. Because the default FRRs were derived from household surveys in SSA countries, they may mis-specify fertility of WLHIV in other regions. Lacking representative measures of HIV prevalence among pregnant women, some Spectrum-AIM users manually adjust the LAF so that modelled estimates of PWLHIV are greater than numbers of PWLHIV receiving ARVs from programme data. In non-SSA regions, more than half of all countries with results published by UNAIDS in 2023 (n=52/83) manually changed the LAF.^10,11^

Large adjustments to the default fertility patterns or uncharacteristically large differences between related model outputs could indicate inaccurate model estimates, inaccurate input data, or inappropriate interpretations of data sources. To identify possible challenges, we used UNAIDS estimates from Spectrum-AIM published in 2023 to establish patterns for HIV prevalence and treatment coverage among pregnant women for each region. We also proposed an algorithm to guide Spectrum-AIM users to critically review the consistency of programmatic estimates with regional patterns and scrutinize surveillance data, programmatic input, and model assumptions.^10,11^ As a case study, we applied the approach and described its impact on estimates of HIV prevalence among pregnant women using Spectrum-AIM estimates for Burkina Faso.

## Methods

We used Spectrum-AIM files submitted by HIV estimation teams to UNAIDS for publication in 2023 to calculate three ratios: (1) HIV prevalence among pregnant women to HIV prevalence among all women 15-49y (‘prevalence ratio’), (2) ART coverage among PWLHIV before the current pregnancy to ART coverage among WLHIV 15-49y (’ART coverage pre-pregnancy ratio’), and (3) the ratio of ART coverage in PWLHIV at delivery to ART coverage in WLHIV 15-49y (‘PMTCT coverage ratio’). We reported these ratios for the year 2022, as they did not vary substantially over the past decade. We summarised ranges for each ratio by UNAIDS region. Values outside of these ranges may indicate inconsistent data for the number of pregnant women on ART, number of PWLHIV, or low predictive power of the local HIV surveillance data to estimate HIV prevalence in the general population.

### Data sources

We extracted outputs from 154 publicly available national or subnational publicly available Spectrum-AIM HIV estimates files submitted by 126 countries and published by UNAIDS in 2023.^11^ Three SSA countries had subnational files (Kenya, Zimbabwe, Ethiopia); the remaining 123 files represented national HIV epidemics created by country teams and submitted to UNAIDS for review and publication. Spectrum-AIM methods are described elsewhere.^19–21^ Briefly, in countries with high HIV prevalence (n= 64, mostly in SSA, including Burkina Faso), adult HIV prevalence and incidence trends were estimated from data on HIV prevalence among (1) nationally-representative samples of adults from household-based surveys and (2) pregnant women attending ANC, sampled in periodic sentinel surveillance up to the mid-2010s and more recently from ANC-RT data.^5^ In countries with lower HIV prevalence, epidemic trends were fit to national HIV and/or AIDS case reports and AIDS-related deaths reported through vital registration with the CSAVR or ECDC models (n= 46) or from HIV prevalence survey and surveillance data among risk groups using the EPP concentrated (n= 27) or AEM models (n= 12).^6,7^ All incidence models accounted for the effects of ART on survival and transmission. Locations and the estimation method for each are reported in Supplementary Table S1.

Spectrum-AIM calculates HIV prevalence among pregnant women from age-specific HIV prevalence of all women 15-49y, age-specific fertility rates,^8^ and HIV fertility rate ratios.^9^ Data on HIV FRRs outside SSA are limited and therefore default HIV FRR patterns for SSA are applied to all regions, despite very different risk populations and contraceptive use. We extracted Spectrum-AIM’s estimates for HIV prevalence among women 15-49y, HIV prevalence among pregnant women, ART coverage among WLHIV 15-49y, and initiation timing for PWLHIV on ART for 2022. We compared HIV prevalence among pregnant women and women 15-49y to nationally representative surveys where available (Supplementary Table S2 and Supplementary Figure S1).

### HIV prevalence, ART, and PMTCT ratios

We calculated the ‘prevalence ratio’ for all Spectrum-AIM files by dividing the HIV prevalence among pregnant women by the HIV prevalence among women 15-49y (equation 1):

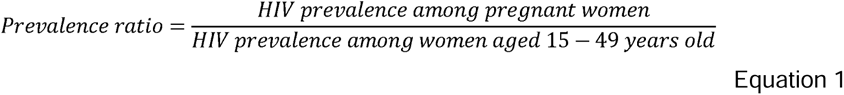

The ‘ART coverage pre-pregnancy ratio’ was calculated as the proportion of PWLHIV on ART before the current pregnancy from programmatic input and Spectrum-AIM’s estimate of PWLHIV, divided by ART coverage among WLHIV 15-49y (equation 2):

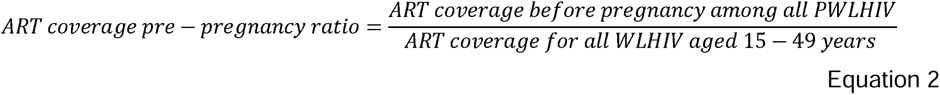

The ‘PMTCT coverage ratio’ was the ratio of total ART coverage (i.e., proportion of PWLHIV who received ART during the pregnancy, started before or during the current pregnancy) and ART coverage for WLHIV 15-49y (equation 3):^24^

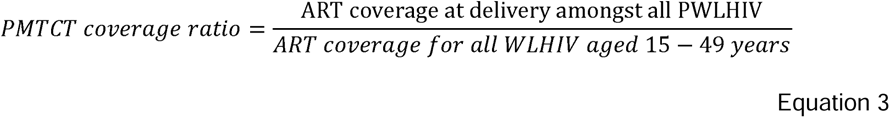

### Identifying typical ranges and outliers by region

We calculated the mean of each ratio by UNAIDS-defined regions, broadly representing variations in epidemic type: eastern and southern Africa (ESA), western and central Africa (WCA), Latin America and Caribbean (LAC), Asia and Pacific (AP), eastern Europe and central Asia (EECA), western and central Europe and North America (WCENA), middle East and northern Africa (MENA). For ESA and WCA, we considered location-specific ratios between 0.75 times lower to 1.25 times higher than the regional mean as ‘typical’ and those outside this range to be outliers. For other regions, we present these ranges but emphasize the influence of local HIV surveillance data on estimating HIV prevalence among pregnant women. Alternate methods to define typical ranges were considered (Supplementary Text S1).

For countries where transmission primarily occurs outside of key populations, we expected prevalence ratios to be less than one (i.e., lower HIV prevalence among pregnant women than all women 15-49y),^25,26^ and the ART pre-pregnancy ratios (on ART before pregnancy) to be less than one, because PWLHIV are on average younger and so acquired HIV more recently.^27,28^ We expected PMTCT coverage ratios to be above one in countries with high ANC-RT coverage and ART for PMTCT uptake, as pregnant women who were untreated before pregnancy should be diagnosed and initiated on ART through ANC-RT.^29^ Outside of SSA, where HIV transmission is mostly among key populations and their partners, and fertility and contracepting patterns are different from SSA, there was little *a priori* information about expected typical relationships for the prevalence, ART pre-pregnancy, or PMTCT ratios.

We further hypothesized that countries with outlier prevalence, ART coverage, or PMTCT coverage ratios had atypical LAFs that were fitted or modified to reconcile discrepant data on HIV prevalence among pregnant women (whether fit to ANC-RT data or estimated by Spectrum-AIM) and number of PWLHIV on ART from programmes. Alternatively, if the distribution of LAFs across countries within a region systematically differed from one, this may indicate that the regional HIV-related FRR parameters in Spectrum-AIM are mis-specified.

To test these hypotheses, we calculated an average LAF by region, weighted according to how close location-specific prevalence, ART pre-pregnancy, and PMTCT ratio (Ratio_L_) were to the regional mean ratio (Ratio_R_). The inverse favours values that are closest to the regional mean ratio. Weighted regional LAFs were calculated separately for all ratios.

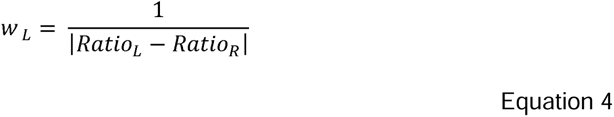

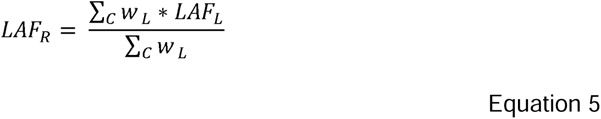

Lastly, we identified inconsistent ART coverage among WLHIV 15-49y and PWLHIV. Inconsistent coverage estimates consisted of: (1) more women receiving ART than estimated WLHIV and (2) large year-to-year differences in the number of women receiving ART. The first indicated inaccurate programme data or modelled output depending on the location, the second indicated possible data recording issues and programmatic changes that should be verified by those familiar with local context.

### Guidance for reviewing data and model outputs with Burkina Faso case study

From the outlier analysis, we developed a six-step algorithm to assess data and model assumptions regarding HIV prevalence and ART coverage among pregnant women (Figure 1). We applied the algorithm using a preliminary Spectrum-AIM file produced by Burkina Faso (WCA region) for the 2023 round of UNAIDS published estimates. This file represents the most up-to-date surveillance and treatment data from the national HIV programme and estimates.

**Figure 1.**
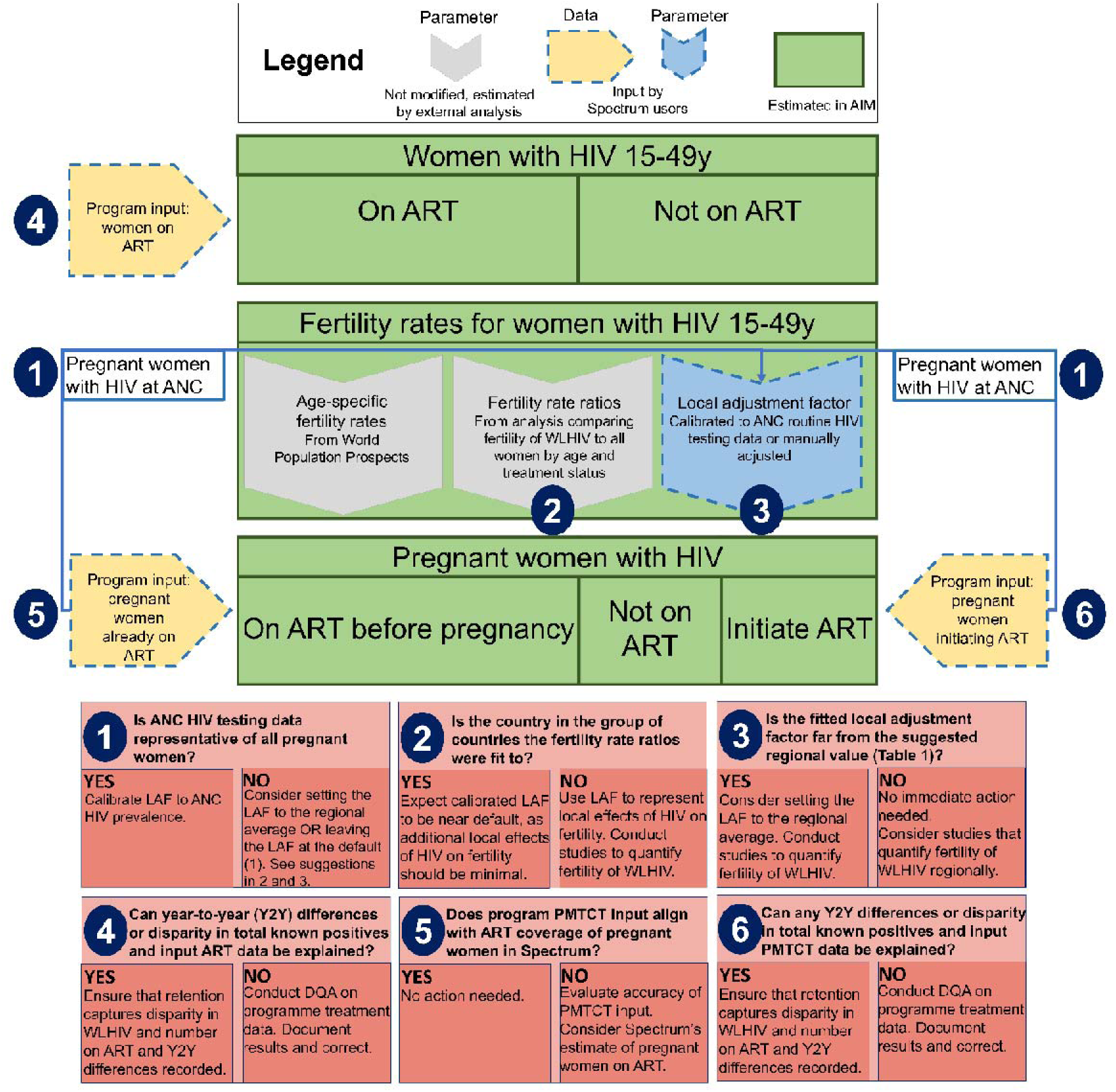
Recommendations to align Spectrum-AIM estimates of HIV prevalence and treatment coverage of pregnant women with regional means.

This study was reviewed and approved by the Research Governance and Integrity team of Imperial College London (ICREC #6300528).

## Results

### Prevalence, ART pre-pregnancy, and PMTCT ratios

In 2022, the mean prevalence ratio was less than one in all regions, ranging from 0.68 in the MENA region to 0.95 in the LAC region (Table 1), indicating that pregnant women typically had lower HIV prevalence than women 15-49y. The ESA and WCA regions had mean prevalence ratios of 0.72 and 0.83, respectively (Table 1). WCA had a higher proportion of outliers than ESA (Figure 2; ESA: 6/46; WCA: 5/25). The regional mean ART coverage pre-pregnancy ratio was lowest in WCA at 0.40, and less than one for all regions, except WCENA and EECA where it was 1.22 and 1.06, respectively (Table 1). Twenty-two countries had ART coverage pre-pregnancy ratios above one (Figure 3). The mean ART coverage pre-pregnancy ratio across all locations was 0.83 and the mean PMTCT coverage ratio was 1.21, and in all regions the mean PMTCT coverage ratio exceeded the ART coverage pre-pregnancy ratio. For all regions except WCA (0.85), the PMTCT coverage ratio exceeded one.

**Figure 2.**
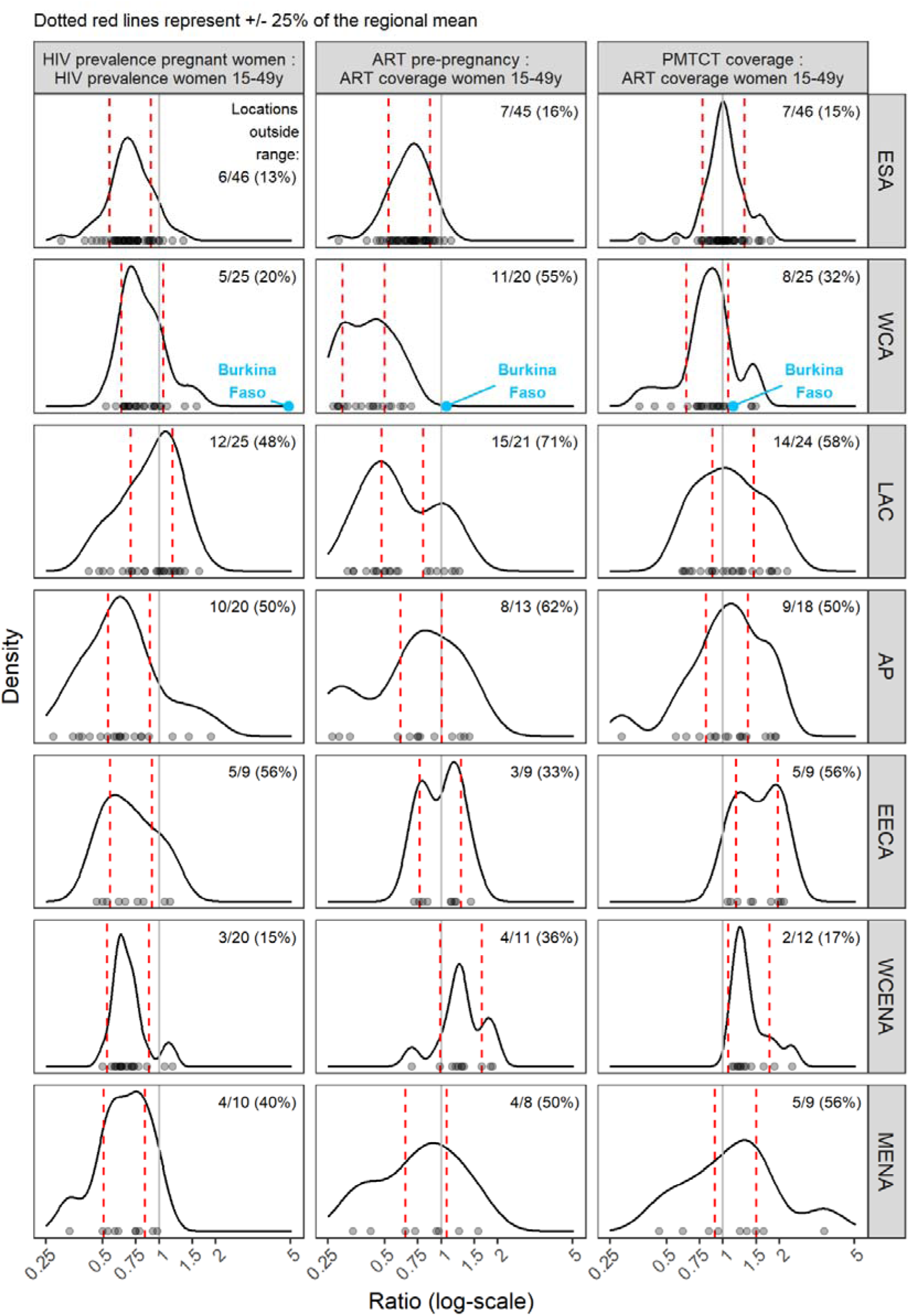
Prevalence, ART pre-pregnancy, and PMTCT ratio by region. Points on the x-axis represent distribution of ratios for all locations. Red dashed lines represent typical ranges of each ratio by region calculated as 0.75 – 1.25 times the mean value. Curved lines show density of ratios by region, with more common ratios represented as peaks. Multimodal distributions represent within region heterogeneity of ratios, meaning the ‘typical’ ranges should be interpreted with caution.

**Figure 3.**
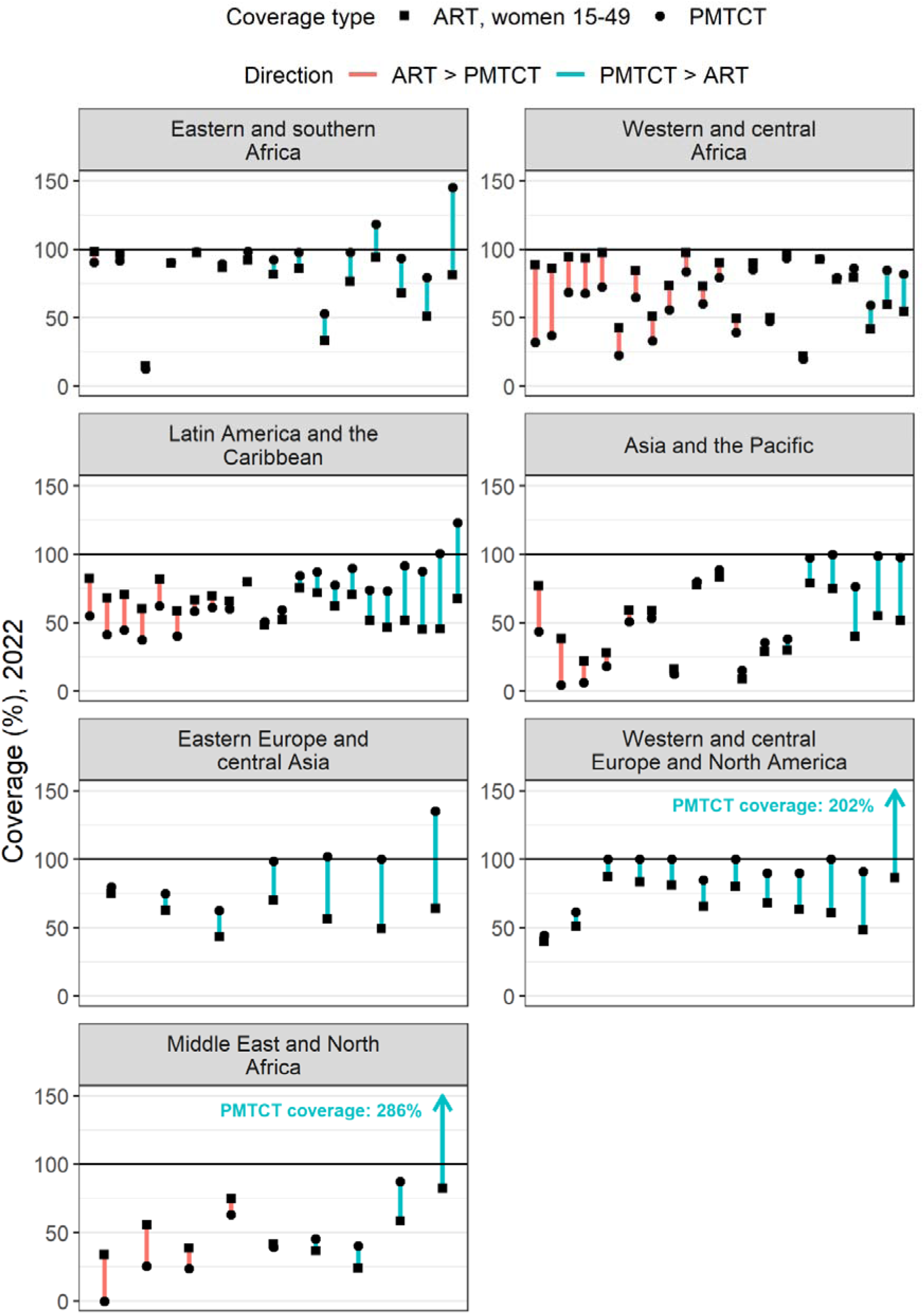
Difference between ART coverage among WLHIV 15-49y and ART coverage at delivery in 2022 for all countries analysed (grouped by region). Length of line indicates size of difference for a given country, colour of line indicates whether ART coverage or PMTCT coverage was higher, and shape of dot indicates the type of coverage.

**Table 1.** Typical ranges of prevalence ratios (defined as HIV prevalence among pregnant women to HIV prevalence among women 15-49y), ART pre-pregnancy (defined as ART coverage among PWLHIV at first ANC to ART coverage among WLIHV 15-49y), and PMTCT (defined as ART coverage started before or during the pregnancy among PWLHIV to ART coverage among WLHIV 15-49y) ratios

| Ratio | UNAIDS Region | Mean<br>(Standard<br>Deviation) | Typical<br>range<br>(0.75 - 1.25x<br>mean) | Typical local<br>adjustment<br>factor<sup>1</sup> |
| --- | --- | --- | --- | --- |
| **Prevalence** | Eastern and southern Africa | 0.72 (0.20) | (0.54 - 0.90) | 1.02 |
|  | Western and central Africa | 0.83 (0.24) | (0.62 - 1.04) | 1.12 |
|  | Latin America and Caribbean | 0.95 (0.29) | (0.71 - 1.18) | 1.62<sup>2</sup> |
|  | Asia and Pacific | 0.73 (0.41) | (0.55 - 0.92) | 1.07<sup>2</sup> |
|  | Eastern Europe and central Asia | 0.73 (0.24) | (0.55 - 0.91) | 1.28<sup>2</sup> |
|  | Western and central Europe and North America | 0.70 (0.17) | (0.53 - 0.88) | 1.07<sup>2</sup> |
|  | Middle East and northern Africa | 0.68 (0.21) | (0.51 - 0.84) | 1.08<sup>2</sup> |
| **ART pre-pregnancy coverage** | Eastern and southern Africa | 0.69 (0.17) | (0.52 - 0.87) | 0.99 |
|  | Western and central Africa | 0.40 (0.16) | (0.30 - 0.50) | 1.27 |
|  | Latin America and Caribbean | 0.67 (0.32) | (0.50 - 0.84) | 1.69<sup>2</sup> |
|  | Asia and Pacific | 0.81 (0.36) | (0.61 - 1.01) | 1.21<sup>2</sup> |
|  | Eastern Europe and central Asia | 1.06 (0.25) | (0.79 - 1.32) | 1.29<sup>2</sup> |
|  | Western and central Europe and North America | 1.22 (0.42) | (0.91 - 1.52) | 1.08<sup>2</sup> |
|  | Middle East and northern Africa | 0.84 (0.45) | (0.63 - 1.05) | 1.37<sup>2</sup> |
| **PMTCT coverage** | Eastern and southern Africa | 1.04 (0.26) | (0.78 - 1.30) | 1.02 |
|  | Western and central Africa | 0.85 (0.32) | (0.64 - 1.06) | 1.19 |
|  | Latin America and Caribbean | 1.19 (0.48) | (0.89 - 1.48) | 2.14<sup>2</sup> |
|  | Asia and Pacific | 1.08 (0.55) | (0.81 - 1.35) | 1.05<sup>2</sup> |
|  | Eastern Europe and central Asia | 1.51 (0.40) | (1.13 - 1.89) | 1.61<sup>2</sup> |
|  | Western and central Europe and North America | 1.47 (0.49) | (1.10 - 1.84) | 1.16<sup>2</sup> |
|  | Middle East and northern Africa | 1.34 (0.95) | (1.00 - 1.67) | 1.20<sup>2</sup> |

<sup>1</sup>Method for calculating detailed in section S1 and Table S4 of SDC
<sup>2</sup>Not calibrated to routine HIV testing done at ANC data

### Atypical ratios within regions

All regions had countries with prevalence ratios outside of the regional typical range (Table 1). Globally, 19 of 154 locations (11 national locations, 8 subnational locations across Ethiopia, Kenya, and Zimbabwe) reported more PWLHIV receiving ART than the estimated number of PWLHIV in 2022. Of these 19 locations, two countries reported more pregnant women on ART before the current pregnancy than total estimated PWLHIV. Every region had countries with ART pre-pregnancy ratios outside of 0.75 – 1.25 times the mean ART pre-pregnancy ratio regional range (Figure 2).

### Burkina Faso case study

We applied the data review steps described in Figure 1 to the Burkina Faso Spectrum-AIM file. Burkina Faso calibrated HIV prevalence among pregnant women to HIV prevalence from ANC-RT data. Prevalence from routine ANC testing in 2022 was 1.19%; Spectrum estimated HIV prevalence among all women 15-49y as 0.7% (0.6 – 0.9%). This differed from the typical relationship where prevalence among pregnant women was less than HIV prevalence among all women.^10^ Step one of the process (Figure 1) revealed that the prevalence ratio in Burkina Faso (1.56) exceeded the typical range for WCA (0.62-1.04, Figure 2). Step two identified Burkina Faso’s LAF parameter of 2.15 was greater than the typical LAF in the region (1.12, Table 1). The third step situated Burkina Faso among countries with a national household survey that informed FRR parameter estimates in Spectrum-AIM, indicating that the default FRR parameters should represent the relative fertility of WLHIV in Burkina Faso. Because both the prevalence ratio and LAF were higher than typical for WCA, we determined that the ANC-RT reported HIV prevalence was higher than expected for a country in WCA.

The Burkina Faso ratios for ART coverage pre-pregnancy (0.47) and PMTCT coverage (1.00) were both within typical ranges for WCA (0.30 - 0.50 for ART pre-pregnancy and 0.64 - 1.06 for PMTCT ratios in WCA, Table 1). We did not find inconsistencies in ART coverage among WLHIV 15-49y in step four. The steps to detect inconsistencies in PMTCT programme data inputs (five and six) indicated the number of WLHIV on PMTCT exceeded the modelled number of PWLHIV in each year between 2015 and 2020 (Figure 4). The discrepancy between pregnant women reported on ART at delivery and estimated number PWLHIV was largest in 2019, when the reported number of PWLHIV on ART at delivery treatment was 1.67 times Spectrum-AIM’s estimate of PWLHIV. Between 2014 and 2015, the reported number of WLHIV receiving PMTCT tripled from 4,285 to 12,937.

**Figure 4.**
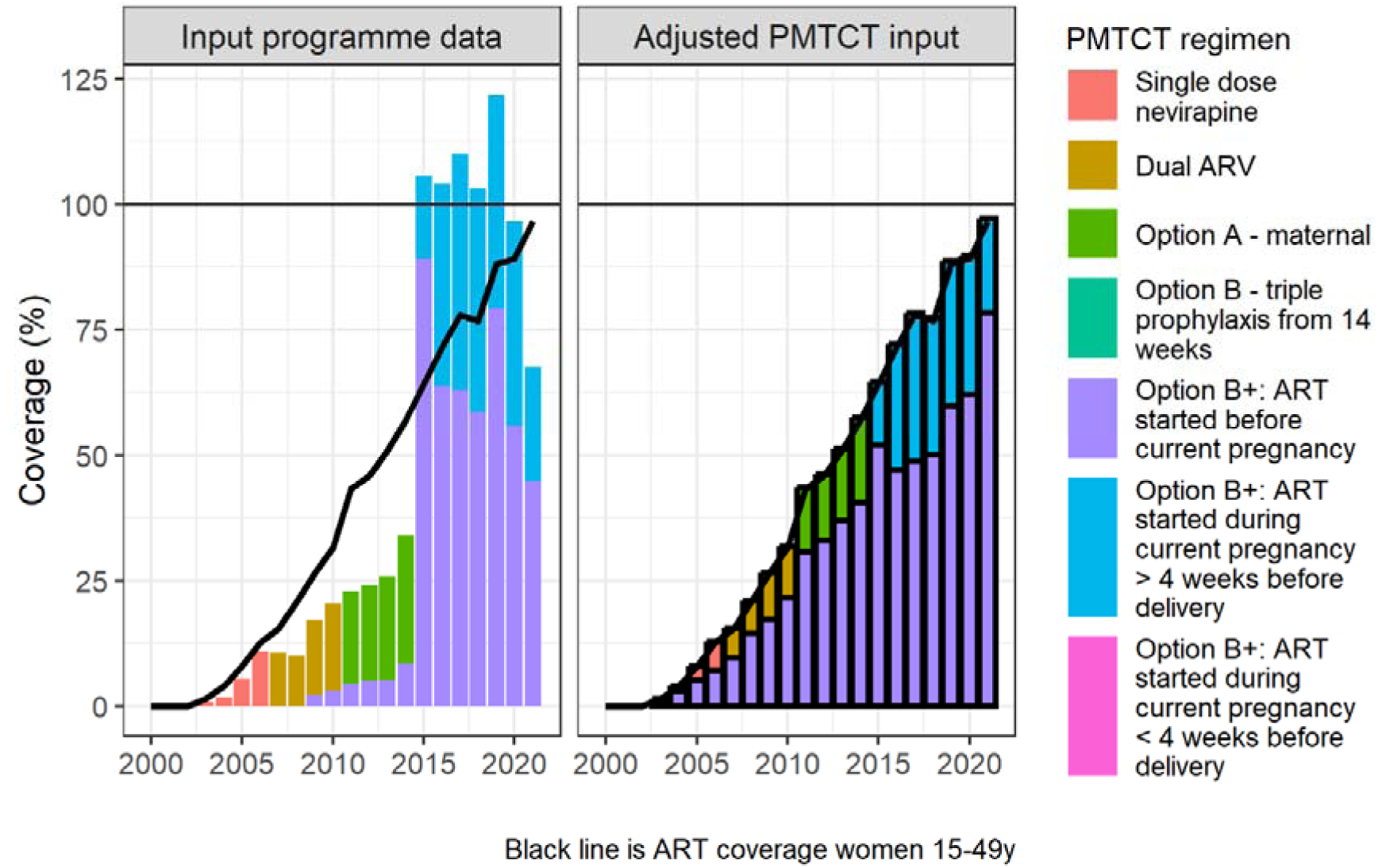
Adjustment of PMTCT programme data for Burkina Faso, shown as the implied percentage coverage relative to Spectrum-AIM-estimated PWLHIV. Bars show estimated PMTCT coverage by regimen; and the line shows ART coverage among all WLHIV 15-49y for comparison. Left: using original PMTCT programme data; right: adjustment to address outcomes of the alignment assessment (Figure 1), as described in section S2 of SDC, bold outlining indicates a modelled value.

The steps outlined in Figure 1 suggested two potential adjustments to reconcile Burkina Faso’s estimates of HIV prevalence among pregnant women: (1) HIV prevalence among pregnant women compared to all women 15-49y was higher than typical for countries in WCA and (2) the number of PWLHIV receiving treatment was inconsistent with the estimated PWLHIV. Both discrepancies could be explained by the inaccurately large number of reported PWLHIV on ART before current pregnancy, which could occur if some PWLHIV on ART were double counted. Per the guidance in steps one and two, we decreased Burkina Faso’s LAF from 2.15 (fit to ANC-RT data) to 1.12 (WCA regional estimate), as an initial step to align Burkina Faso’s prevalence ratio with regional estimates. During steps three and four, we did not change the FRR or ART coverage among women 15-49y, since Burkina Faso’s FRR was fit to national household survey data, and we did not identify inconsistencies in ART coverage among women 15-49y. Because the PMTCT ratio was within the range of typical values for WCA (Table 1), we applied the reported distribution of ART initiation timings annually to ART coverage among women 15-49y to produce new to the overall level of PMTCT coverage (Figure 4, and Supplementary Text S2). Reducing the LAF and updating the number of women receiving antiretrovirals for PMTCT resulted in a new prevalence ratio of 1.07, consistent with other countries in the region.

## Discussion

Our study assessed estimates of HIV prevalence and treatment coverage among pregnant women compared to all adult women by global regions. We developed a data review process for Spectrum-AIM users to assess the relationship between ANC and ART related programme data inputs and assumptions about fertility among WLHIV. This algorithm relies on three ratios (prevalence, ART coverage pre-pregnancy, and PMTCT coverage), prompting users to interrogate discrepancies between programme and surveillance data and Spectrum-AIM assumptions and to align estimates of HIV prevalence among pregnant women and ART coverage at delivery with other locations in the region.

HIV prevalence among pregnant women in 2022 was lower than among all women 15-49y in all regions, typically by 20-30%. Prevalence ratios showed less variation across countries in regions with larger HIV epidemics compared to regions with transmission more concentrated among key populations and their partners. The greater variation in the latter is likely due to diverse epidemiological factors and variations in surveillance data reliability and estimation approaches. There are three notable differences in the respective groups’ Spectrum-AIM models and input data: (1) most countries in SSA use the same type of data (HIV testing from ANC and representative household surveys) to fit overall adult HIV prevalence, whereas concentrated epidemic countries use surveillance among key populations (primarily non-ANC) data, (2) for countries that do use ANC data outside of SSA, HIV testing at ANC is often targeted at women at higher risk to HIV,^30,31^ and (3) default FRRs were derived from household survey data from SSA and may be more applicable there.

In most regions, ART coverage before pregnancy was lower than ART coverage among all adult WLHIV, but by varying degrees (regional means ranging 0.40 – 0.87, excluding EECA and WCENA). ART coverage before pregnancy was higher in EECA (1.06) and WCENA (1.22). All countries in EECA and WCENA with ART pre-pregnancy ratios greater than one were estimated with the CSAVR model, which uses national HIV diagnoses and/or AIDS case reports and AIDS-related deaths reported through vital registration to estimate age-specific HIV prevalence (Supplementary Table S1).^6^ Assessing typical results for the prevalence, ART coverage pre-pregnancy, and PMTCT ratios for countries without data that directly measures HIV prevalence among pregnant women requires more nuance than for countries with ANC-RT data.

Interpreting outlier ratios involves scrutinizing both local HIV surveillance data for total population prevalence estimation and its implications for estimating HIV prevalence among pregnant women.

1. Countries should assess the reliability of data used to estimate age-specific HIV prevalence among adults, considering documented biases in HIV surveillance data.^32–35^ Where death registration data are used, countries should consider how frequent misclassification of cause of death is.^35^
2. If data on PMTCT provision is reliable, countries could consider manual adjustments to the LAF to align the number of PWLHIV with the reported number of PWLHIV on ART from programmes. More work should be done to estimate the relative fertility of WLHIV in that epidemic setting rather than using the FRR estimated from SSA household surveys.

While the proposed ratios describe relationships between surveillance data, programmatic input, and model assumptions, they are intrinsically linked through the model and cannot be evaluated independently. Aligning one ratio with the mean regional value may cause another ratio to diverge from the regional value; balancing these ratios requires an iterative process of assessing the accuracy of different input data sources and making justified adjustments. While there is no gold standard source to compare these ratios to, using ratios calculated from external sources (such as PHIA and/or DHS surveys) in conjunction with regional ranges to identify outliers is a starting point to scrutinizing implausible model inputs.

The Burkina Faso case study illustrates this. Burkina Faso’s prevalence ratio was higher than many locations in the WCA region accompanied by a high LAF fit to national ANC-RT data. The atypically high prevalence ratio suggested that Burkina Faso’s ANC-RT data overstated HIV prevalence among pregnant women relative to all women 15-49y. Previous studies found ANC testing clinics in Burkina Faso disproportionately serve urban areas with higher HIV prevalence among pregnant women than national HIV prevalence among pregnant women.^36^ The difference in clientele is corroborated by lower access to multiple ANC visits before delivery in rural areas of Burkina Faso.^36,37^ Ultimately, the Burkina Faso estimation team determined the programme data were not robust for use in HIV estimation indicated by low concordance between prevalence values from routine programme data and sentinel surveillance data.

We suggest Spectrum-AIM users complete this review during the annual UNAIDS-supported HIV estimation process. Regions with epidemics primarily among key populations will have more heterogeneous fertility among WLHIV and access to care than countries with transmission among the heterosexual population.^39–42^ In this case, adjustments based on regional trends may be less appropriate and Spectrum-AIM users should prioritize insights about the local epidemic context and ANC coverage data to inform the more accurate estimates of HIV prevalence and ART coverage among pregnant women. Additionally, countries should triangulate results with additional valid data sources, including testing of HIV exposed infants with mother infant pairing.^43–45^ Future work should examine patterns across epidemic types and the relationship between pregnant women and key population HIV prevalence to improve internal consistency in Spectrum-AIM’s estimates of HIV prevalence among pregnant women. Investing in validation with external data not input to the Spectrum-AIM model will produce more accurate estimates of HIV prevalence among pregnant women and number of vertical HIV transmissions. Such data triangulation is well illustrated by Sibanda *et al.*, who integrated data from four sources (including Spectrum-AIM results) to identify gaps in the PMTCT care cascade in Zimbabwe, improving programme results.^46^

Our work provides recommendations and a standardized approach to examining and, where needed, improving estimates of HIV prevalence among pregnant women within the Spectrum-AIM modelling process. We provide a path forward for targeted data collection and quality review for further refining these parameterizations. Future work could expand this analysis to include metrics measured through other data systems, such as early infant diagnosis and vital registration. Improvements in estimates of the number of pregnant women with HIV will aid measuring and addressing PMTCT needs, thereby decreasing instances of vertical transmission, towards the elimination of maternal to child transmission.

## Conflicts of Interest and Source of Funding

The authors declare no conflicts of interest. This research was supported by the National Institute of Allergy and Infectious Diseases of the National Institutes of Health under award number 1R01AI152721-01A1, UNAIDS, and the MRC Centre for Global Infectious Disease Analysis (reference MR/R015600/1), jointly funded by the UK Medical Research Council (MRC) and the UK Foreign, Commonwealth & Development Office (FCDO), under the MRC/FCDO Concordat agreement and is also part of the EDCTP2 programme supported by the European Union.

A preliminary analysis of this work was virtually presented in October 2022 to the UNAIDS Reference Group on HIV Estimates, Modelling, and Projections. For open access, the author has applied a Creative Commons Attribution (CC BY) license to any Author Accepted Manuscript version arising.

## Data Availability

All data produced in the present study are available upon reasonable request to the authors

## Notes

### Competing Interest Statement

The authors have declared no competing interest.

### Author Declarations

Ethics committee/IRB of Imperial College London gave ethical approval for this work

