## supplemental material for "Guidance for triangulating data and estimates of HIV prevalence among pregnant women and coverage of PMTCT using the Spectrum AIDS Impact Module"

Supplemental Digital Content

Magdalene K. Walters^1§^, Eline L. Korenromp^2^, Anna Yakusik^2^, Ian Wanyeki^2^, André Kaboré^3,^ Arthur Poimouribou^4^, Célestine Ki^4^, Coumbo Dao^5^, Paul Bambara^4^, Salam Derme^4^, Théophile Ouedraogo^4^, Kai Hon Tang^1^, Marie-Claude Boily^1^, Mary Mahy^2^, Jeffrey W. Imai-Eaton^1,7^

^1^ MRC Centre for Global Infectious Disease Analysis, School of Public Health, Imperial College London, London, United Kingdom

^2^ Data for Impact Department, Joint United Nations Programme on HIV/AIDS, Geneva, Switzerland

^3^ Data for Impact Division, Joint United Nations Programme on HIV/AIDS, Ouagadougou, Burkina Faso

^4^ Permanent Secretary of the National Council for the Fight against AIDS and Communicable Infections, Burkina Faso

^5^ Direction de la Santé de la Famille (DSF), Ouagadougou, Burkina Faso

^7^ Center for Communicable Disease Dynamics, Department of Epidemiology, Harvard T.H. Chan School of Public Health, Boston, MA, USA

Table of Contents

Table S1	3-5

Table S2	6

Table S3	7

Figure S1	8

Figure S2	8

Figure S3	9

S1. Weighting method to derive regional LAF values: sensitivity	10-11

Table S4	10

Table S5	11

S2. Burkina Faso treatment modelling  	11

Table S6	11

**Table S1.** UNAIDS Spectrum files in each region and incidence estimation model used

| Region | Location | Incidence model used in 2023-round Spectrum-AIM estimate |
| --- | --- | --- |
| Asia and the Pacific | Afghanistan | EPP, concentrated |
|  | Australia | ECDC |
|  | Bangladesh | AEM |
|  | Bhutan | EPP, concentrated |
|  | Cambodia | AEM |
|  | Fiji | EPP, concentrated |
|  | Indonesia | AEM |
|  | Lao People’s Democratic Republic | AEM |
|  | Malaysia | AEM |
|  | Maldives | EPP, concentrated |
|  | Mongolia | AEM |
|  | Myanmar | AEM |
|  | Nepal | AEM |
|  | New Zealand | CSAVR |
|  | Pakistan | AEM |
|  | Papua New Guinea | EPP, generalized |
|  | Philippines | AEM |
|  | Sri Lanka | AEM |
|  | Thailand | AEM |
|  | Timor-Leste | EPP, concentrated |
| Eastern Europe and central Asia | Albania | CSAVR |
|  | Armenia | CSAVR |
|  | Belarus | EPP, concentrated |
|  | Georgia | CSAVR |
|  | Kyrgyzstan | CSAVR |
|  | Montenegro | CSAVR |
|  | Republic of Moldova, Left bank | EPP, concentrated |
|  | Republic of Moldova, Right bank | EPP, concentrated |
|  | Tajikistan | CSAVR |
| Eastern and southern Africa | Angola | EPP, generalized |
|  | Botswana | EPP, generalized |
|  | Comoros | EPP, concentrated |
|  | Eritrea | EPP, generalized |
|  | Ethiopia, Addis Abeba | EPP, generalized |
|  | Ethiopia, Affar | EPP, generalized |
|  | Ethiopia, Amhara | EPP, generalized |
|  | Ethiopia, Benis Gumz | EPP, generalized |
|  | Ethiopia, Dire Dawa | EPP, generalized |
|  | Ethiopia, Gambela | EPP, generalized |
|  | Ethiopia, Harari | EPP, generalized |
|  | Ethiopia, Oromiya | EPP, generalized |
|  | Ethiopia, Sidama | EPP, generalized |
|  | Ethiopia, SNNP | EPP, generalized |
|  | Ethiopia, Somali | EPP, generalized |
|  | Ethiopia, Tigray | EPP, generalized |
|  | Kenya, Central | EPP, generalized |
|  | Kenya, Coast | EPP, generalized |
|  | Kenya, Eastern | EPP, generalized |
|  | Kenya, Nairobi | EPP, generalized |
|  | Kenya, North Eastern | EPP, generalized |
|  | Kenya, Nyanza | EPP, generalized |
|  | Kenya, Rift Valley | EPP, generalized |
|  | Kenya, Western | EPP, generalized |
|  | Lesotho | EPP, generalized |
| Eastern and southern Africa (continued) | Madagascar | EPP, concentrated |
|  | Malawi | EPP, generalized |
|  | Mozambique | EPP, generalized |
|  | Namibia | EPP, generalized |
|  | Rwanda | EPP, generalized |
|  | South Africa | Direct incidence input |
|  | South Sudan | EPP, generalized |
|  | Swaziland | EPP, generalized |
|  | Uganda | EPP, generalized |
|  | United Republic of Tanzania | EPP, generalized |
|  | Zambia | EPP, generalized |
|  | Zimbabwe, Bulawayo | EPP, generalized |
|  | Zimbabwe, Harare Chitungwiza | EPP, generalized |
|  | Zimbabwe, Manicaland | EPP, generalized |
|  | Zimbabwe, Mashonaland Central | EPP, generalized |
|  | Zimbabwe, Mashonaland East | EPP, generalized |
|  | Zimbabwe, Mashonaland West | EPP, generalized |
|  | Zimbabwe, Masvingo | EPP, generalized |
|  | Zimbabwe, Matabeleland North | EPP, generalized |
|  | Zimbabwe, Matabeleland South | EPP, generalized |
|  | Zimbabwe, Midlands | EPP, generalized |
| Latin America and Caribbean | Argentina | CSAVR |
|  | Bahamas | CSAVR |
|  | Belize | CSAVR |
|  | Bolivia | EPP, concentrated |
|  | Brazil | Direct incidence input |
|  | Colombia | EPP, concentrated |
|  | Costa Rica | CSAVR |
|  | Cuba | CSAVR |
|  | Dominican Republic | EPP, concentrated |
|  | Ecuador | EPP, concentrated |
|  | El Salvador | CSAVR |
|  | Grenada | CSAVR |
|  | Guatemala | EPP, concentrated |
|  | Guyana | EPP, concentrated |
|  | Haiti | EPP, generalized |
|  | Honduras | EPP, concentrated |
|  | Mexico | CSAVR |
|  | Nicaragua | EPP, concentrated |
|  | Panama | CSAVR |
|  | Paraguay | CSAVR |
|  | Peru | EPP, concentrated |
|  | St. Kitts | CSAVR |
|  | Trinidad and Tobago | EPP, concentrated |
|  | Uruguay | CSAVR |
|  | Venezuela | CSAVR |
| Middle East and North Africa | Algeria | CSAVR |
|  | Jordan | CSAVR |
|  | Kuwait | CSAVR |
|  | Lebanon | CSAVR |
|  | Libyan Arab Jamahiriya | CSAVR |
|  | Morocco | EPP |
|  | Qatar | CSAVR |
|  | Syrian Arab Republic | CSAVR |
|  | Tunisia | EPP |
|  | Yemen | CSAVR |
| Western and central Africa | Benin | EPP, generalized |
|  | Burkina Faso | EPP, generalized |
|  | Burundi | EPP, generalized |
|  | Cameroon | EPP, generalized |
|  | Cape Verde | EPP, concentrated |
|  | Central African Republic | EPP, generalized |
|  | Chad | EPP, generalized |
|  | Congo | EPP, generalized |
|  | Côte d’Ivoire | EPP, generalized |
|  | Democratic Republic of the Congo | EPP, generalized |
|  | Equatorial Guinea | EPP, generalized |
|  | Gabon | EPP, generalized |
|  | Gambia | EPP, generalized |
|  | Ghana | EPP, generalized |
|  | Guinea | EPP, generalized |
|  | Guinea-Bissau | EPP, generalized |
|  | Liberia | EPP, generalized |
|  | Mali | EPP, generalized |
|  | Mauritania | EPP, concentrated |
|  | Niger | EPP, concentrated |
|  | Senegal | EPP, concentrated |
|  | Sierra Leone | EPP, generalized |
|  | São Tomé and Príncipe | EPP, concentrated |
|  | Togo | EPP, generalized |
| Western and central Europe and North America | Canada | Direct incidence input |
|  | Croatia | CSAVR |
|  | Cyprus | ECDC |
|  | Czech Republic | CSAVR |
|  | Denmark | CSAVR |
|  | France | Direct incidence input |
|  | Greece | ECDC |
|  | Iceland | CSAVR |
|  | Ireland | CSAVR |
|  | Italy | CSAVR |
|  | Lithuania | Direct incidence input |
|  | Luxembourg | CSAVR |
|  | Malta | CSAVR |
|  | Netherlands | ECDC |
|  | Portugal | CSAVR |
|  | Romania | CSAVR |
|  | Serbia | CSAVR |
|  | Slovakia | CSAVR |
|  | Slovenia | ECDC |
|  | TFYR Macedonia | CSAVR |

**Table S2.** Data source descriptions and assessment criteria

| Source | Household surveys (DHS and PHIA) | Modelled results (Spectrum/ AIM) | |
| --- | --- | --- | --- |
|  | HIV prevalence | HIV prevalence | ART coverage |
| Population and representativeness | Adults 15-49y  Nationally representative | Nationally or sub-nationally representative | Nationally or sub-nationally representative |
| Data collection method | HIV testing | Fit to HIV prevalence from household surveys, time trend fit to ANC HIV testing | Numerator reported by country teams  Denominator estimated from number PLHIV |
| Data type | HIV-focused, cross-sectional, household-based, nationally representative surveys of children and adults | National or subnational results for all years 1990-2022 at 5-year age groups | National or subnational results for all years 1990-2022 at 5-year age groups |
| Potential issues | Potential non-response bias^32,47^ | Relationship between general population and ANC HIV prevalence varies by epidemic type and level of ANC attendance^26^ | Number on ART may exceed estimated PLHIV  Potential inaccuracies in data reporting (e.g. year to year inconsistencies or large increases or decreases in coverage)^48^ |

**Table S3**. Prevalence ratios from most recent DHS or PHIA survey

| Country | Year | Survey | Prevalence pregnant women | Prevalence women 15-49y | Prevalence ratio |
| --- | --- | --- | --- | --- | --- |
| Angola | 2015 | DHS | 1.3% | 2.6% | 0.51 |
| Burkina Faso | 2010 | DHS | 0.6% | 1.2% | 0.48 |
| Burundi | 2016 | DHS | 0.7% | 1.2% | 0.57 |
| Cameroon | 2018 | DHS | 3.4% | 3.4% | 0.98 |
| Chad | 2014 | DHS | 1.2% | 1.8% | 0.66 |
| Congo | 2009 | DHS | 3.7% | 4.1% | 0.91 |
| Congo Democratic Republic | 2013 | DHS | 0.6% | 1.6% | 0.37 |
| Côte d'Ivoire | 2012 | DHS | 2.7% | 4.6% | 0.58 |
| Eswatini | 2016 | PHIA | 35.2% | 27.0% | 1.31 |
| Ethiopia | 2017 | PHIA | 1.8% | 3.0% | 0.58 |
| Gabon | 2012 | DHS | 5.2% | 5.8% | 0.89 |
| Gambia | 2013 | DHS | 1.4% | 2.1% | 0.65 |
| Ghana | 2014 | DHS | 2.8% | 2.8% | 0.97 |
| Guinea | 2018 | DHS | 1.7% | 1.6% | 1.03 |
| Kenya | 2018 | PHIA | 4.2% | 4.9% | 0.86 |
| Lesotho | 2016 | PHIA | 24.7% | 25.6% | 0.97 |
| Liberia | 2013 | DHS | 4.6% | 2.4% | 1.92 |
| Malawi | 2016 | PHIA | 8.3% | 10.6% | 0.78 |
| Mali | 2012 | DHS | 1.9% | 1.3% | 1.40 |
| Mozambique | 2015 | DHS | 10.1% | 15.4% | 0.65 |
| Namibia | 2017 | PHIA | 11.9% | 12.6% | 0.94 |
| Niger | 2012 | DHS | 0.2% | 0.4% | 0.57 |
| Rwanda | 2018 | PHIA | 2.4% | 3.0% | 0.81 |
| São Tomé and Príncipe | 2008 | DHS | 1.5% | 1.3% | 1.18 |
| Senegal | 2017 | DHS | 0.4% | 0.5% | 0.86 |
| Sierra Leone | 2013 | DHS | 1.1% | 1.7% | 0.67 |
| South Africa | 2016 | DHS | 29.1% | 27.7% | 1.05 |
| Tanzania | 2012 | DHS | 3.2% | 6.2% | 0.53 |
| Togo | 2013 | DHS | 1.6% | 3.1% | 0.52 |
| Uganda | 2017 | PHIA | 5.6% | 6.2% | 0.89 |
| Zambia | 2018 | DHS | 10.4% | 14.3% | 0.73 |
| Zimbabwe | 2016 | PHIA | 11.1% | 13.6% | 0.82 |

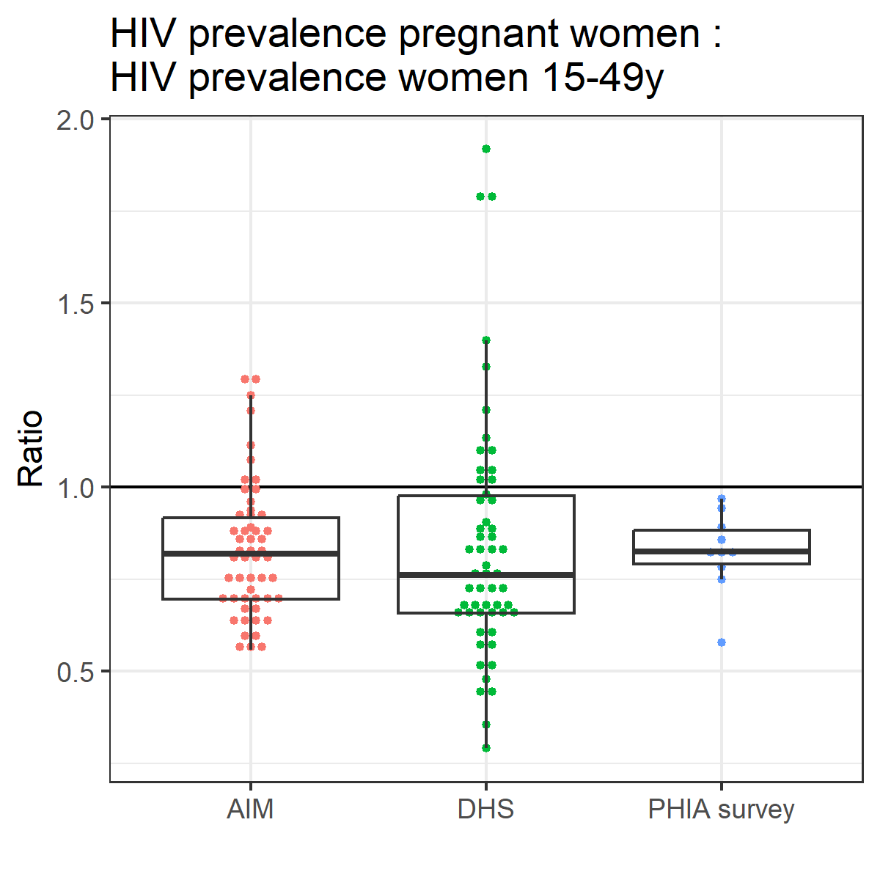

**Figure S1.** Prevalence ratios for all location-years with DHS or PHIA surveys and Spectrum-AIM estimates for the corresponding location-years

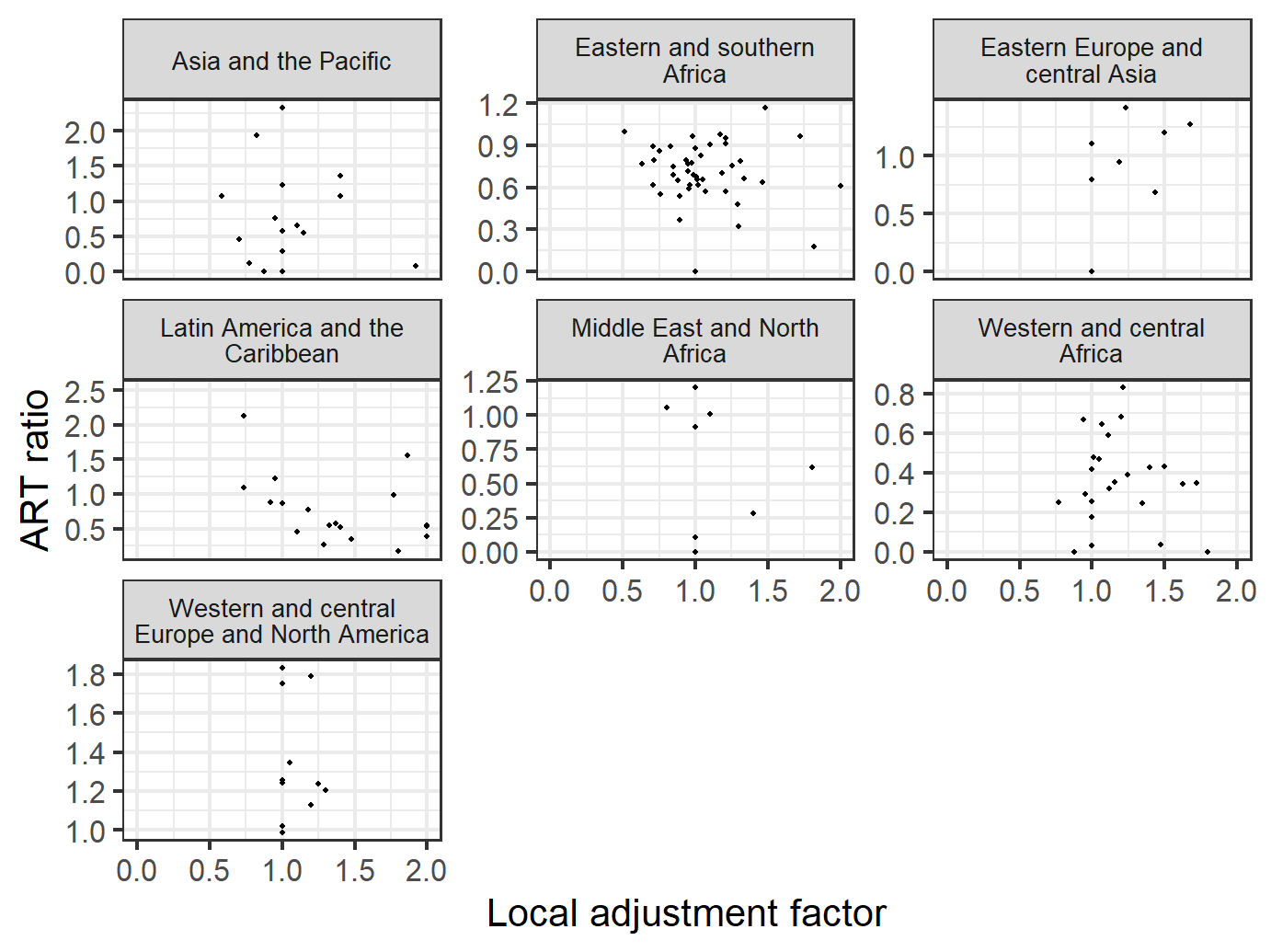

**Figure S2.** Local adjustment factor by region and relationship with ART pre-pregnancy ratio

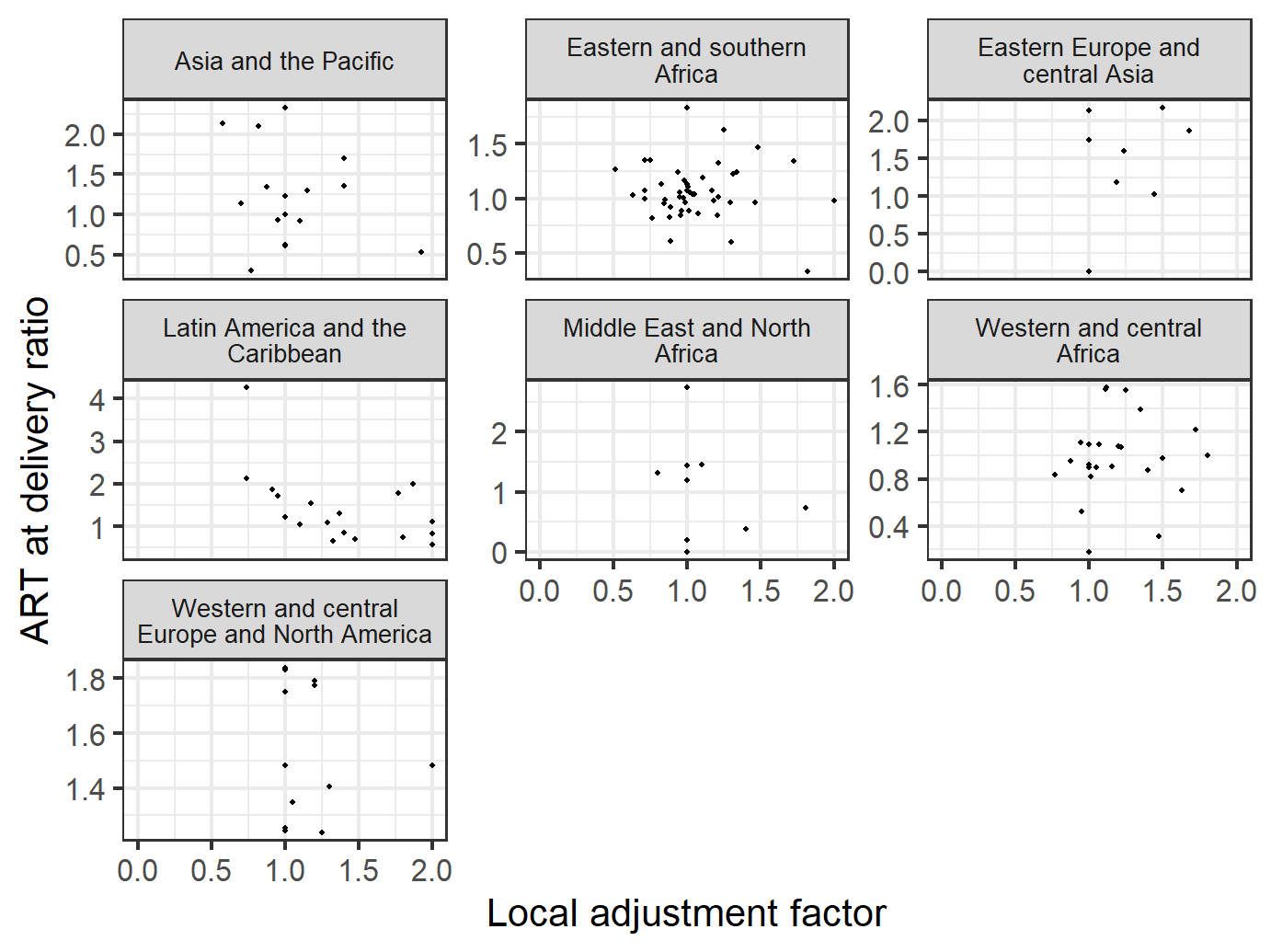

**Figure S3.** Local adjustment factor by region and relationship with PMTCT ratio.

### S1. Weighting method to derive regional LAF values: sensitivity analysis

To provide regionally typical ranges for values of LAF that produce prevalence, ART, and PMTCT ratios, we calculated a weighted mean of LAFs from 2023 Spectrum-AIM files, across all locations within a region. The weights prioritized LAFs in countries with prevalence/ART pre-pregnancy/PMTCT ratios closer to the regional mean. We considered alternate methods to provide these benchmarks, as a sensitivity analysis for this weighting method.

When comparing different methods of creating regional mean LAFs, we examined both how many countries fell outside the range of regional values, and the difference in LAFs across metrics. In the table below we present various methods to calculate regional ranges for prevalence/ART pre-pregnancy/PMTCT ratios and countries falling outside each range (Table S4) and regional LAFs (Table S5).

We present this just for the prevalence ratio for simplicity; the same was applied for the other two ratios (not shown) but patterns were similar. For the prevalence ratio, these comparisons show that broadly, the regional prevalence ratios are robust against alternative calculations of the typical value. The counts of outliers are also robust, except for WCENA region, which had very low within region heterogeneity (Figure 1, main text).

**Table S4.** Regional ranges of prevalence ratios, compared between alternative across outliering method

| Region | Mean approach^1^ | | Median approach | | IQR approach | |
| --- | --- | --- | --- | --- | --- | --- |
|  | Mean  (range) | Outliers | Median (range) | Outliers | Median (range) | Outliers |
| Asia and Pacific | 0.73  (0.55 – 0.92) | 10/20 | 0.62  (0.46 – 0.77) | 11/20 | 0.62  (0.47 – 0.80) | 10/20 |
| Eastern Europe and central Asia | 0.73  (0.55 – 0.91) | 5/9 | 0.66  (0.49 – 0.82) | 4/9 | 0.66  (0.53 – 0.83) | 4/9 |
| Eastern and southern Africa | 0.72  (0.54 – 0.90) | 6/46 | 0.69  (0.52 – 0.87) | 8/46 | 0.69  (0.61 – 0.82) | 12/46 |
| Latin America and Caribbean | 0.95  (0.71 – 1.18) | 12/25 | 0.98  (0.74 – 1.23) | 12/25 | 0.98  (0.71 – 1.16) | 12/25 |
| Middle East and northern Africa | 0.68  (0.51 – 0.84) | 4/10 | 0.68  (0.51 – 0.85) | 4/10 | 0.68  (0.55 – 0.78) | 6/10 |
| Western and central Africa | 0.83  (0.62 – 1.04) | 5/25 | 0.76  (0.57 – 0.95) | 7/25 | 0.76  (0.69 – 0.95) | 12/25 |
| Western and central Europe and North America | 0.70  (0.53 – 0.88) | 3/20 | 0.65  (0.49 – 0.81) | 3/20 | 0.65  (0.61 – 0.73) | 10/20 |

^1^Used in final analysis

We also considered the extent to which the regional LAF was sensitive to its calculation method. In Table S5 three options for calculating a regional LAF: weighted (as described in the main text and listed in Table 1), unweighted, and median. To calculate the unweighted mean, we simply averaged all LAFs within a region. To calculate the median, we found the median value of the LAF within a region.

**Table S5.** Averaging method for LAF selection

| Region | Weighted mean^1^ | Unweighted mean | Median |
| --- | --- | --- | --- |
| Asia and Pacific | 1.05 | 1.14 | 1.00 |
| Caribbean | 1.45 | 1.84 | 1.35 |
| Eastern Europe and central Asia | 1.20 | 1.36 | 1.35 |
| Eastern and Southern Africa | 0.94 | 1.04 | 1.00 |
| Latin America | 1.68 | 1.73 | 1.86 |
| Middle East and Northern Africa | 1.03 | 1.15 | 1.00 |
| Western and central Africa | 1.02 | 1.04 | 1.00 |
| Western and central Europe and North America | 1.01 | 1.13 | 1.00 |

^1^Used in final analysis

The regional LAF was sensitive to the calculation method. The unweighted mean made the regional LAF more prone to being skewed by outliers, while the median increased the relevance of countries which do not adjust their LAF. This was as expected and validates our choice of using the weighted mean to calculate the regional LAF.

### S2. Burkina Faso treatment modelling

Because Burkina Faso seemed to have an unreasonable increase the number of women on PMTCT between 2014 and 2015, we replaced this input with extrapolated values. To do this, we used the *PMTCT ratio* to scale the overall PMTCT coverage and then applied the existing proportions of each treatment type of overall PMTCT coverage to get the coverage of each option for all years between 2000 and 2022. The resulting treatment coverage by PMTCT option for the year 2022 is listed in Table S6 below.

**Table S6.** Burkina Faso treatment modelling

| PMTCT option | 2022 input to AIM (#) | Proportion of total | 2022 ART coverage  15-49y (%) | WCA  ART at delivery coverage :  ART coverage 15-49y | 2022 suggested value (%) |
| --- | --- | --- | --- | --- | --- |
| Single dose nevirapine | 0 | 0 | 98.20% | 0.85 | 0 |
| Dual ARV | 0 | 0 |  |  | 0 |
| Option A- maternal | 0 | 0 |  |  | 0 |
| Option B- triple prophylaxis from 14 weeks | 0 | 0 |  |  | 0 |
| Option B+: ART started before current pregnancy | 2454 | 0.51 |  |  | 42.20% |
| Option B+: ART started during current pregnancy > 4 weeks before delivery | 2402 | 0.49 |  |  | 41.30% |
| Option B+: ART started during current pregnancy < 4 weeks before delivery | 0 | 0 |  |  | 0 |
